# Predicting Inpatient Risk of Mortality in Diabetic Patients Using Administrative Data and Machine Learning: An External Validation Study Using SPARCS

**DOI:** 10.1101/2025.06.05.25329076

**Authors:** Ali Mirza, Tobechi Nwokeji

## Abstract

**Objectives:** To evaluate whether machine learning models trained solely on administrative and demographic data can predict inpatient APR Risk of Mortality in diabetic patients.

**Design:** Retrospective cohort study using New York State SPARCS data from 2021 and 2022.

**Setting:** New York Statewide Planning and Research Cooperative System (SPARCS) data from 2021 and 2022.

**Participants:** Adult inpatient admissions (age ≥18) with a diagnosis of diabetes mellitus.

**Primary outcome measure:** APR-DRG Risk of Mortality (ROM), classified as Minor, Moderate, Major, or Extreme.

**Results:** XGBoost outperformed logistic regression and random forest across all metrics. On the 2022 validation set, XGBoost achieved the highest accuracy (46.5%), macro AUC (0.699), weighted F1-score (0.458), and the lowest Brier score for the Extreme class (0.052). SHAP analysis identified length of stay, age group, and payer type as key predictors.

**Conclusions:** Even without clinical data, administrative features contain non-random signals relevant for mortality risk stratification. These models, especially XGBoost, may help hospitals flag high-risk patients early using routinely available data, aiding triage and planning before labs or vitals are available.

**Strengths and limitations of this study:**

- This study is one of the first to apply machine learning to publicly available SPARCS data to predict APR-DRG Risk of Mortality in diabetic inpatients.
- We evaluated three models using temporally distinct training and validation cohorts, simulating real-world model deployment across calendar years.
- Model interpretability was addressed using SHAP, providing transparent insights into feature contributions and enabling clinician-facing explanation.
- The models relied solely on administrative and demographic data, limiting predictive fidelity due to the absence of clinical features such as laboratory values or vital signs.
- Risk of Mortality labels were derived from APR-DRG software and may be influenced by coding practices rather than objective clinical outcomes.

## Introduction

Inpatient mortality risk prediction plays a central role in hospital triage, resource allocation, and early clinical decision-making. Accurate identification of high-risk patients can inform care escalation, monitoring protocols, and discharge planning. While modern risk scores such as the All-Patient Refined Diagnosis Related Groups (APR-DRG) Risk of Mortality (ROM) index are widely used in hospital operations, their underlying methodologies are proprietary, and their reliance on fully coded clinical data limits their utility early in the patient admission.

In parallel, machine learning (ML) methods have demonstrated strong predictive capabilities across diverse clinical outcomes, but many require granular electronic health record (EHR) data such as lab values, vital signs, or imaging-derived variables. These features may not be immediately available at the time of admission, particularly in fragmented or resource-limited health systems.

To address this gap, we investigated whether ML models trained on administrative and demographic features alone––without any labs, vitals, or physician-entered clinical notes—could effectively predict APR-DRG Risk of Mortality. We focused on diabetic patients, a population at heightened risk for adverse inpatient outcomes. Using publicly available discharge-level administrative data from the New York Statewide Planning and Research Cooperative System (SPARCS), we trained and externally validated three ML models: Random Forest, XGBoost, and Logistic Regression. Models were trained on 2021 data and validated on the 2022 data to simulate temporal generalizability.

Our goal was not to replicate proprietary ROM scores exactly, but rather to evaluate how much mortality risk signal is embedded in basic administrative features alone. We compared models using multiple performance metrics including accuracy, macro-averaged AUC, weighted F1, Brier score, and quadratic-weighted Cohen’s Kappa. We also explored model calibration and feature attribution using SHAP values to support interpretability.

This study offers a practical baseline for administrative-only mortality prediction and contributes to the broader conversation about transparent, interpretable ML tools in clinical operations.

## Methods

### Study Design and Data Source

This was a retrospective cohort study conducted using the New York Statewide Planning and Research Cooperative System (SPARCS), a publicly available all-payer discharge database that captures hospital-level administrative data for all inpatient encounters in New York State. We used two pre-cleaned datasets derived from SPARCS for the years 2021 and 2022. Each dataset included only adult patients (aged 18 years and older) with an inpatient admission and a diagnosis of diabetes mellitus, as coded by ICD-10 or CCS categories. Patients with missing target values were excluded from the analysis.

The 2021 dataset was used for model training and internal cross-validation, while the 2022 dataset served as an independent external validation cohort. There was no temporal overlap between cohorts. This study followed the Transparent Reporting of a multivariable prediction model for Individual Prognosis Or Diagnosis (TRIPOD) guidelines for prognostic model development and evaluation.

### Outcome Variable

The primary outcome was the All Patient Refined Diagnosis Related Groups (APR-DRG) Risk of Mortality (ROM), a categorical variable computed by proprietary software and widely used for benchmarking risk adjustment in hospital administrative data. The ROM score consists of four levels: Minor, Moderate, Major, and Extreme. For modeling purposes, these labels were numerically encoded as 0, 1, 2, and 3, respectively. The variable was treated as a multiclass classification target rather than an ordinal or regression outcome.

### Predictor Variables and Preprocessing

Each record in the datasets contained nine categorical predictor variables. These included age group, three-digit ZIP code prefix, gender, race, ethnicity, primary payer type, categorized length of stay, admission type, and clinical classification software (CCS) diagnosis group. These variables were selected to reflect information typically available at the point of admission and intentionally excluded clinical measurements such as laboratory results, vital signs, or imaging findings.

All predictor variables were one-hot encoded prior to model training. Feature columns in the 2022 validation set were reindexed to match the training feature space derived from the 2021 cohort. Any feature levels present in the training set but absent in the validation set were imputed with zero values to maintain alignment in dimensionality.

The training cohort (2021) included 20,806 patient admissions, and the external validation cohort (2022) included 20,270 admissions. Table 1 presents the class distribution of the outcome variable in both cohorts.

**Table 1.** Distribution of APR-DRG Risk of Mortality Classes in Training and Validation Cohorts.

| **Risk of Mortality Class** | **Training Cohort (2021), n (%)** | **Validation Cohort (2022), n (%)** |
| --- | --- | --- |
| Minor | 6,302 (30.3%) | 5,842 (28.8%) |
| Moderate | 7,416 (35.6%) | 7,075 (34.9%) |
| Major | 6,034 (29.0%) | 6,168 (30.4%) |
| Extreme | 1,054 (5.1%) | 1,185 (5.8%) |
| **Total** | **20,806 (100%)** | **20,270 (100%)** |

### Model Development

Three supervised classification models were developed: multinomial logistic regression with L2 regularisation, random forest with balanced class weights, and XGBoost using the multi:softprob objective. All models were implemented using standard Python libraries (`scikit-learn` and `xgboost`) with default hyperparameters unless otherwise specified.

Each model was trained on the full 2021 training dataset. To assess internal stability, 5-fold stratified cross-validation was conducted on the training data using weighted F1 score as the primary evaluation metric. After cross-validation, each model was refit on the entire 2021 dataset and evaluated on the 2022 external validation set.

### Model Evaluation

Models were evaluated on the external validation set using multiple performance metrics. These included overall accuracy, weighted F1 score, quadratic-weighted Cohen’s kappa, macro-averaged area under the receiver operating characteristic curve (AUC), and Brier score for the Extreme ROM class. The macro-AUC metric treated the multiclass problem as a one-vs-rest structure and was computed by averaging class-specific AUCs.

To statistically compare model performance, McNemar’s test was applied to the prediction outcomes of each pair of models. The test compared disagreement patterns in prediction correctness between two classifiers on a sample-by-sample basis, assessing whether observed differences in prediction accuracy were significant beyond chance.

The final training cohort included 20,806 inpatient admissions, and the external validation cohort included 20,270 admissions. The distribution of risk categories across both cohorts is shown in Table 1.

Given the observed class imbalance—with the “Extreme” risk category comprising approximately 5% of the sample—class weighting was applied during model training to mitigate skewed performance and to improve sensitivity for rare outcomes. Class-specific performance metrics such as Brier score and AUC were reported to reflect the model’s calibration and discrimination, particularly in underrepresented classes.

### Model Explainability

To enhance transparency and interpretability, we applied SHapley Additive exPlanations (SHAP) to the XGBoost model. SHAP values were computed for all samples in the 2022 validation cohort and summarised using a bar plot, which visualised the average magnitude of each feature’s contribution to the model’s predictions. This approach enabled global interpretability of the XGBoost model across all predicted risk categories.

## Results

### Model Performance

Across all evaluation metrics, the XGBoost model consistently outperformed both logistic regression and random forest classifiers on the external validation set from 2022.

Table 2 summarizes the performance of all three models. XGBoost achieved the highest accuracy (46.5%), weighted F1-score (0.458), quadratic-weighted Cohen’s Kappa (0.424), and macro-averaged AUC (0.699). It also achieved the lowest Brier score (0.052) for the “Extreme” risk class, indicating superior calibration for predicting high-severity outcomes.

**Table 2.** Model performance on external validation set (2022)

| Metric | Logistic Regression | Random Forest | XGBoost |
| --- | --- | --- | --- |
| Accuracy | 0.417 | 0.412 | **0.465** |
| F1-score (Weighted) | 0.422 | 0.410 | **0.458** |
| Quadratic-weighted Kappa | 0.420 | 0.321 | **0.424** |
| Macro AUC | 0.698 | 0.640 | **0.699** |
| Brier Score<br>(Extreme class, lower = better) | 0.088 | 0.065 | **0.052** |

*Bold* indicates best performance for each metric.

### Confusion Matrix and ROC Analysis

The confusion matrix for XGBoost (Figure 1) revealed that most model misclassifications occurred between adjacent APR severity categories, particularly between Moderate and Major risk levels.

**Figure 1.**
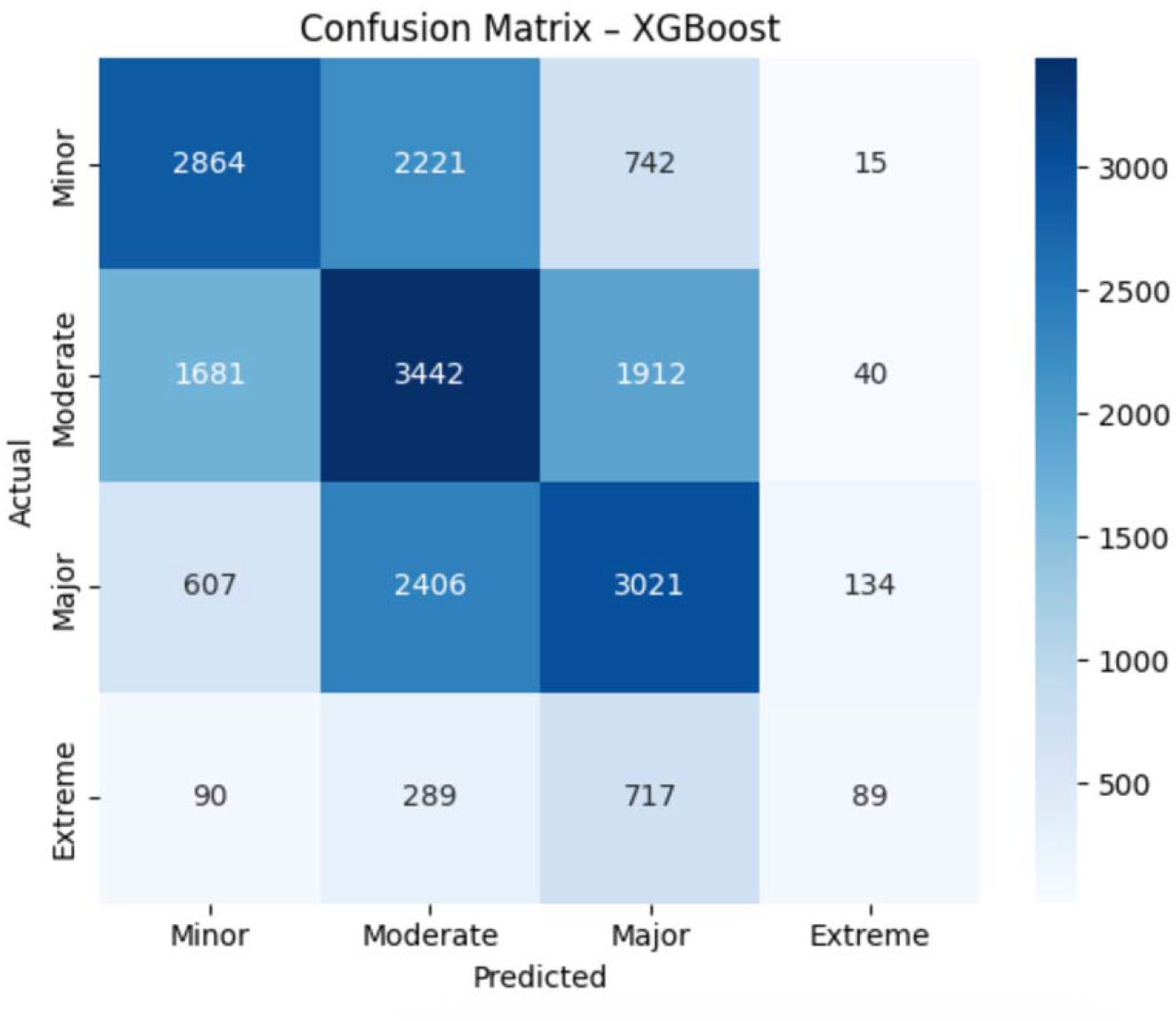
Confusion matrix for XGBoost model on 2022 validation set

Receiver operating characteristic (ROC) curves were plotted for each APR risk class (Figure 2). The model demonstrated reasonable discrimination, with AUCs of 0.75 (Minor), 0.59 (Moderate), 0.70 (Major), and 0.76 (Extreme). The lower AUC for the Moderate class reflects greater overlap in patient features across adjacent categories.

**Figure 2.**
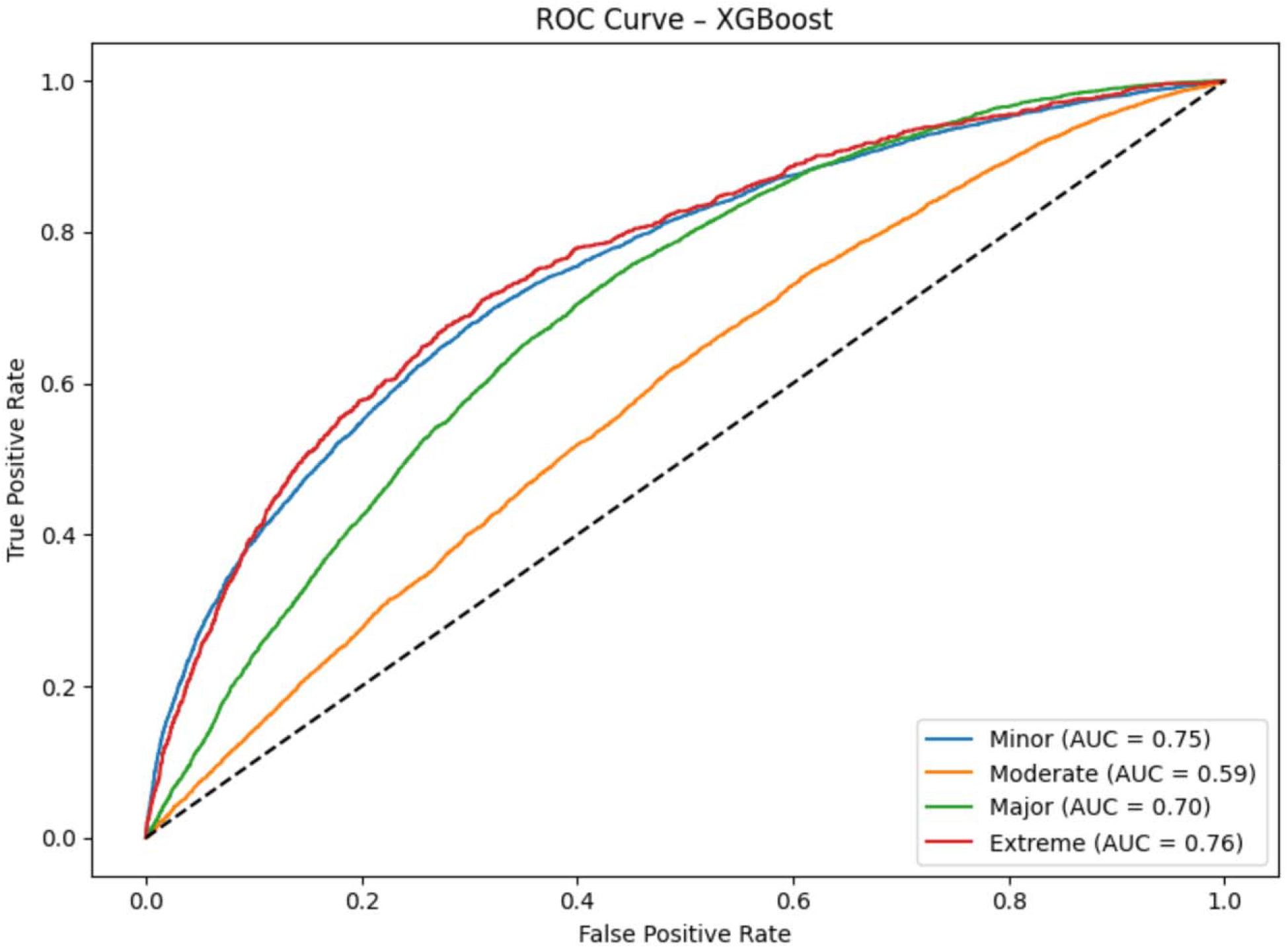
ROC curves by class for XGBoost

### Feature Importance

SHAP analysis was performed to identify the features most influential in the XGBoost model’s predictions (Figure 3). Length of stay variables (particularly for 1–3 days) and young age groups (under 30) had the greatest predictive influence. Payer type and ZIP code were also among the top contributors.

**Figure 3.**
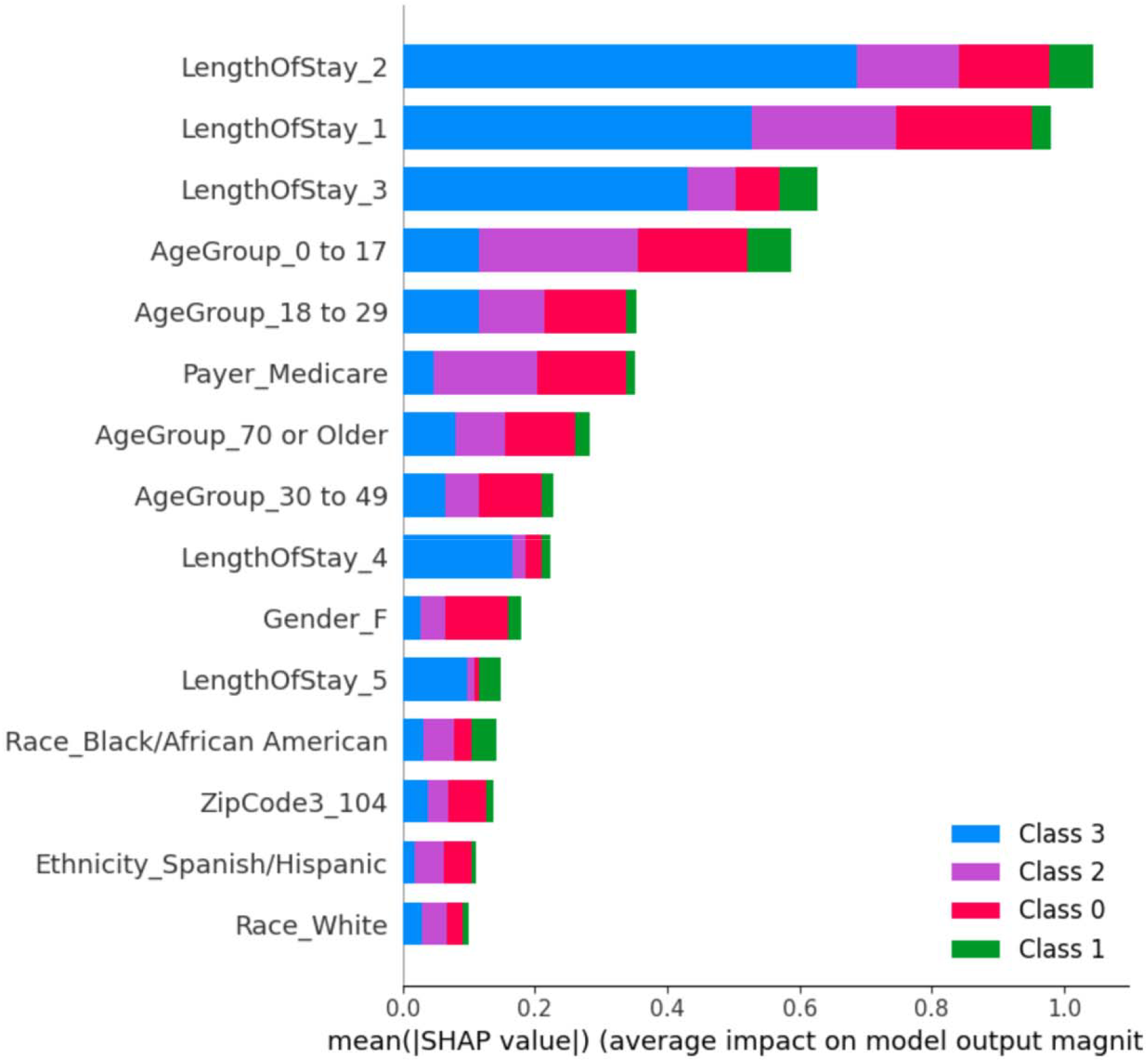
Top 15 most important features for XGBoost (SHAP summary plot)

### Statistical Comparison

McNemar’s test confirmed that the performance difference between XGBoost and both logistic regression (p = 1.9×10□□^3^) and random forest (p = 3.6×10□□□) was statistically significant. This suggests that XGBoost yields meaningfully different and more accurate predictions.

## Discussion

This study demonstrates that machine learning models—particularly XGBoost—can effectively predict APR-DRG Risk of Mortality in diabetic inpatients using routinely collected administrative data. When externally validated on a temporally distinct cohort from 2022, the XGBoost model consistently outperformed both logistic regression and random forest classifiers across all evaluation metrics, including accuracy, macro-averaged AUC, quadratic-weighted Kappa, and Brier score (Table 2).

### Model Performance and Clinical Implications

XGBoost achieved the highest overall accuracy (46.5%) and F1-score (45.8%), as well as the strongest agreement with the ground truth as measured by quadratic-weighted Kappa (0.424). The model also demonstrated superior calibration for the “Extreme” risk class, achieving the lowest Brier score (0.052), suggesting its predictions were not only discriminative but well-calibrated for the highest-severity patients. These findings are particularly relevant in resource-constrained settings where early identification of high-risk patients can guide triage, discharge planning, and clinical escalation.

The relatively low macro-AUC values across all models, even for XGBoost (0.699), reflect the inherent difficulty of this multi-class classification problem, particularly in differentiating between adjacent severity classes. This challenge is further illustrated in the ROC curves (Figure 2), where discrimination for the “Moderate” risk group (AUC = 0.59) was noticeably lower than for “Extreme” (AUC = 0.76) or “Minor” (AUC = 0.75), possibly due to feature overlap and diagnostic ambiguity in the moderate-risk population.

### Feature Importance and Unexpected Predictors

The SHAP summary plot (Figure 3) reveals that the most predictive features for mortality risk classification were length of stay (LOS) and age group. The three most important features were all LOS categories (1, 2, and 3 days), which aligns with prior literature suggesting that atypically short or long hospital stays can signal higher risk trajectories. However, a notable and somewhat unexpected finding was the high predictive contribution of younger age groups, specifically “0 to 17” and “18 to 29.” This warrants attention.

While counterintuitive, this may reflect the relatively low base rate of severe outcomes in younger populations, whereby those who do present with complications may represent disproportionately high-risk cases (e.g., Type 1 diabetes with ketoacidosis or other acute metabolic derangements). The model may be capturing these outlier patterns, which could explain their elevated SHAP values. Alternatively, this pattern may be a function of unmeasured confounding or imbalance in the training data, and warrants further investigation through subgroup analysis or inclusion of interaction terms in future modeling efforts.

Payer type (Medicare) and zip code–derived socioeconomic markers (e.g., ZipCode3_104) also emerged as meaningful predictors. These likely act as proxies for structural determinants of health such as access to care, comorbidity burden, or care fragmentation, further supporting the model’s potential utility in flagging systemic risk not captured by clinical metrics alone.

### Strengths and Limitations

A key strength of this study is the use of temporally external validation, which provides a more rigorous test of model generalizability than cross-validation alone. Additionally, the use of SHAP values provides interpretable insights into feature contributions, which is critical for clinical adoption.

However, several limitations must be acknowledged. First, the use of administrative data, while widely available, lacks granularity regarding laboratory results, vital signs, and clinical documentation that could further improve model accuracy. Second, the APR-DRG Risk of Mortality score itself may be subject to coding variation across institutions. Finally, while XGBoost outperformed traditional models, its overall predictive performance—particularly for intermediate risk classes—remains suboptimal for direct clinical deployment without further refinement.

### Future Directions

Future work should consider integrating richer EHR data sources, exploring longitudinal modeling of risk over time, and validating the approach across institutions and populations. Moreover, qualitative analysis of the high-importance but surprising features (e.g., young age groups) may reveal hidden clinical or sociological phenomena that traditional analyses have missed.

## Conclusion

This study demonstrates the potential of using administrative hospital data and machine learning models, particularly XGBoost, to predict APR-DRG Risk of Mortality among diabetic inpatients. Our findings suggest that XGBoost not only outperforms traditional models such as logistic regression and random forest in discriminative performance but also provides better-calibrated predictions for high-severity cases, as reflected by its lower Brier score for the “Extreme” class.

The integration of SHAP-based model interpretation highlighted clinically intuitive features such as length of stay, while also uncovering less-anticipated contributors, including younger age groups. These results underscore both the promise and complexity of applying machine learning methods to real-world health data, where predictive performance and clinical plausibility must be weighed together.

Despite limitations, the approach shows potential for early risk stratification. Future research should integrate clinical variables and extend to other high-risk populations.

These models may serve as a pragmatic early triage layer prior to clinician review, particularly in environments lacking real-time access to structured electronic health record data.

## Acknowledgements

The authors thank the New York State Department of Health for providing public access to the SPARCS database.

## Funding

This research received no specific grant from any funding agency in the public, commercial or not-for-profit sectors.

## Competing interests

The authors declare no competing interests.

## Patient and public involvement

Patients or the public were not involved in the design, conduct, or reporting of this research.

## Ethics approval

This study used publicly available, de-identified data from the SPARCS database. Ethics approval was not required.

## Data availability statement

All data used in this study are publicly available through the New York State SPARCS database (https://www.health.ny.gov/statistics/sparcs/).

The full codebase for data preprocessing, model training, evaluation, and SHAP-based interpretability is available at: https://github.com/thisisalimirza/diabetesROM_MLModel

## Author contributions

Conceptualization: Ali Mirza, Tobechi Nwokeji

Data curation: Ali Mirza, Tobechi Nwokeji

Methodology: Ali Mirza

Formal analysis: Ali Mirza

Software: Ali Mirza

Writing–original draft: Ali Mirza

Writing–review & editing: Ali Mirza, Tobechi Nwokeji

## Notes

### Competing Interest Statement

The authors have declared no competing interest.

### Funding Statement

This study did not receive any funding.

### Author Declarations

The study used only openly available human data that were publicly accessible before the initiation of the study. The dataset was obtained from the New York Statewide Planning and Research Cooperative System (SPARCS), maintained by the New York State Department of Health. All data are de-identified and publicly available at: https://www.health.ny.gov/statistics/sparcs/

